# Antibody response against SARS-CoV-2 spike protein in people with HIV after COVID-19 vaccination

**DOI:** 10.1101/2024.06.07.24308586

**Authors:** María José Muñoz-Gómez, Pablo Ryan, Marta Quero-Delgado, María Martin-Vicente, Guillermo Cuevas, Jorge Valencia, Eva Jimenez-Gonzalez, Natalia Blanca, Amalia Martinez de Gandara, Gerardo Redondo, Vicente Mas, Mónica Vázquez, Juan Torres-Macho, Isidoro Martinez, Salvador Resino

## Abstract

**Background:** People with HIV (PWH) often have a suboptimal response to vaccines, raising concerns about the efficacy of coronavirus disease 2019 (COVID-19) vaccines in this population. We aimed to evaluate the humoral immune response to the B.1 lineage and Omicron variant in PWH following COVID-19 vaccination.

**Methods:** We conducted a prospective study on 19 PWH who received a two-dose series of the COVID-19 mRNA vaccine and a booster six months later. Participants without HIV infection were included as a healthy control (HC) group. The humoral response to the COVID-19 vaccine (anti-SARS-CoV-2 S IgG levels and ability to block ACE2-S interaction) against both the original B.1 lineage and the Omicron variant was measured by immunoassays.

**Results:** The humoral response in PWH was very strong (GMFR >8) after the second dose and strong (GMFR >4) after the booster dose for both the B.1 lineage and the Omicron variant. We found similar humoral responses to the B.1 lineage and Omicron variant between PWH and HC groups after the second and booster doses (q-value >0.05). The COVID-19 vaccine generated a significantly weaker humoral response against the Omicron variant compared to the B.1 lineage in both groups (q-value <0.05). However, this response improved after the booster dose, although the inhibition of ACE2-S interaction remained weaker in PWH.

**Conclusions:** PWH generated a strong humoral response to the COVID-19 vaccine against both the B.1 lineage and the Omicron variant, similar to individuals without HIV infection. However, our data suggest the need for booster doses to improve immunogenicity and update COVID-19 vaccines for new variants like Omicron.

## Introduction

Human immunodeficiency virus (HIV) infection causes significant alterations in the immune system, particularly in people with HIV (PWH) without antiretroviral therapy (ART). However, even in PWH who have achieved HIV suppression with ART, chronic immune impairment may persist [1], potentially contributing to suboptimal antibody production following vaccination, resulting in diminished vaccine efficacy [2].

PWH are a high-risk group for severe coronavirus disease 2019 (COVID-19), with a 38% higher risk of dying from COVID-19 compared to people without HIV [3]. Consequently, PWH were designated as a priority group for COVID-19 vaccination [2]. The most effective COVID-19 vaccines in preventing severe COVID-19 are the COVID-19 mRNA vaccines, BNT162b2 (Pfizer-BioNTech) and mRNA-1273 (Moderna). However, considering that PWH may exhibit suboptimal responses to other vaccines [4], concerns remain about the potential efficacy of COVID-19 vaccines in this population. Besides, PWH were not included in clinical trials [2], so most of the available information comes from observational studies.

COVID-19 mRNA vaccines induce specific neutralizing antibodies against the spike glycoprotein (S) of Severe Acute Respiratory Syndrome Coronavirus 2 (SARS-CoV-2), which mediates the binding to the angiotensin-converting enzyme 2 (ACE2) receptor on host cells [5, 6]. While numerous studies have established the safety of COVID-19 mRNA vaccines among PWH, there are conflicting data regarding the vaccine-induced humoral immune response in this population [7]. Some studies suggest that there is no difference between PWH and HIV-negative individuals concerning antibody kinetics, peak titers, and neutralization activity [8]. Conversely, other researchers have demonstrated that the antibody response in PWH is correlated with their CD4 cell count levels and viremia, with a compromised immune response in PWH with a CD4 below 200 cells per μL [9, 10].

SARS-CoV-2 mutates quickly, evolving into new variants that increase transmissibility and virulence or reduce the effectiveness of COVID-19 vaccines [2]. The Omicron variant is highly transmissible and resistant to vaccine-induced immunity. Antibody and neutralizing responses against Omicron are significantly lower compared to those against the wild-type SARS-CoV-2 [11, 12]. In the general population, completing a primary vaccination series with two doses offers limited protection against severe Omicron outcomes, but a booster dose increases protection. However, data on PWH are limited.

## Objective

We aimed to evaluate the humoral immune response against the B.1 lineage and Omicron variant generated in response to the COVID-19 vaccine in PWH.

## Materials and Methods

### Study design

We conducted a prospective study on 19 PWH who received the COVID-19 mRNA vaccine, based on the Wuhan-Hu-1 strain, for the first time between February 2021 and July 2021 at the Hospital Universitario Infanta Leonor (HUIL) in Madrid, Spain. We included a healthy control (HC) group of participants without HIV, matched by age and sex to the PWH group.

The vaccination schedule followed international guidelines [13], consisting of two doses of a COVID-19 mRNA vaccine with a 28-day interval, followed by a third booster dose six months later. At the first and second dose (n=44), 18 PWH received the Moderna mRNA-1273 vaccine, one PWH received the Pfizer BNT162b2 mRNA vaccine, and 25 HC received the Moderna mRNA-1273 vaccine. At the booster dose (n=27), 15 PWH received the Moderna mRNA-1273 vaccine, eight HC received the Moderna mRNA-1273 vaccine, and seven HC received the Pfizer BNT162b2 mRNA vaccine.

The HUIL Ethics Committee approved the study (Ref.: 030-21). The research followed the Declaration of Helsinki, and all participants gave their informed consent before enrollment.

### Clinical data and samples

Participant characteristics were collected from the hospital’s electronic medical records. Data were stored using the Research Electronic Data Capture system [14 TN, USA] [14].

Blood samples were collected in ethylene diamine tetra-acetic acid tubes at three points: the first COVID-19 vaccination (baseline), around four weeks after the second vaccine dose, and around ten weeks after the booster dose. All samples were processed and preserved at −80°C in the HUIL until analysis.

### Previous SARS-CoV-2 infection

Participant plasma samples were tested for SARS-CoV-2 infection at baseline, after the second dose, and after the booster dose of the COVID-19 vaccine. The test detected IgG, IgA, and IgM antibodies against SARS-CoV-2 N protein using a commercial enzyme-linked immunosorbent assay (ELISA) (Platelia SARS-CoV-2 Total Ab, Bio-Rad Laboratories Inc., Hercules, California, USA). A sample was considered positive if the optical density ratio between the test and control samples was greater than or equal to 1 (ratio ≥1.0). This cut-off has a sensitivity of 94.7% and a specificity of 97.5% [15].

### Immunoassay for anti-SARS-CoV-2 S IgG quantification

A complete description of the protocols and materials for antibody quantification is provided by Martin-Vicente et al. [16]. We used the plasmid pαH, kindly provided by Dr. McLellan (University of Texas, Austin, USA), encoding the S protein ectodomain (residues 1-1208) of SARS-CoV-2 2019-nCOV (GenBank: MN908947). Two HexaPro constructs were created by mutagenesis to stabilize the prefusion spike protein. For the first construct (B.1 linage), the following substitutions were made: glycine at residue 614 (D614G), a “GSAS” substitution at the furin cleavage site (residues 682–685), and proline at residues 817, 892, 899, 942, 986, and 987. For the second construct (SARS-CoV-2 S Omicron, B.1.1.529), the natural cleavage site “RRAR” (residues 682-685) was included, along with these Omicron specific mutations: A67V, Δ69-70, T95I, G142D/Δ143-145, Δ211/L212I, ins214EPE, G339D, S371L, S373P, S375F, K417N, N440K, G446S, S477N, T478K, E484A, Q493R, G496S, Q498R, N501Y, Y505H, T547K, D614G, H655Y, N679K, P681H, N764K, D796Y, N856K, Q954H, N969K, and L981F).

The plasmid encoding the ACE2 SARS-CoV-2 cell receptor (residues 1-165) was also constructed and fused to a StrepTag.

Antibodies against the S protein were titrated using an ELISA assay. Serial 1:3 dilutions of plasma samples (starting from 1:50 and ending at 1:36450), were incubated with the purified S protein ectodomain. One phase exponential decay least-squares fit curves, and the area under the curve was calculated using GraphPad Prism 9.0.

Antibody inhibition of the ACE2-S protein interaction was tested by ELISA. Serum samples were diluted serially 1:2, starting from 1:10 to 1:320. These dilutions were incubated with the S protein before adding the ACE2 receptor complexed with StrepTactin-peroxidase. A 2016 sera pool from people who tested negative for anti-S antibodies was used as a control. After background subtraction, the inhibition percentage was calculated as [1 - (OD_493_ test plasma / OD_493_ control plasma)] × 100 %.

### Statistical analysis

IBM SPSS Statistics 25.0 (SPSS INC, Armonk, NY, USA) and Stata 15.0 (StataCorp, Texas, USA) were used for statistical analysis. GraphPad Prism 9.0 (GraphPad Software, Inc., San Diego, CA, USA) was used to create the figures. The significance level was set at p<0.05 (two-tailed).

The two main factors analyzed were the study population (PWH vs. HC) and the immune response to SARS-CoV-2 variants (B.1 vs. Omicron). The primary outcome was the humoral response to the COVID-19 vaccine, measured by anti-SARS-CoV-2 S IgG levels and their ability to block ACE2-S interaction against both the original B.1 lineage and the Omicron variant after the second dose and booster dose. Humoral response data were log10-transformed.

Generalized linear mixed models (GLMM) were used to calculate the geometric mean fold rise (GMFR) and their 95% CI between post- and pre-vaccination. The interpretation of the GMFR values was as follows: GMFR > 8 indicated a very strong humoral response with a significant amount of antibodies, GMFR 4 – 8 indicated a strong humoral response, and GMFR <4 indicated a weak humoral response.

GLMM also evaluated the impact of two main factors (study population and SARS-CoV-2 variants) on the COVID-19 vaccine humoral response, providing differences among groups. P-values were corrected by the False Discovery Rate (q-value). The rate of non-responders (AUC = 0) was compared using the Chi-squared test.

## Results

### Patient characteristics

Baseline characteristics of PWH are shown in **Table 1**. All patients were on ART, two had a detectable viral load (>50 HIV RNA copies/mL), and six had a CD4 count below 200 cells/mm^3^.

**Table 1.** Characteristics of people with HIV infection at baseline. **Statistics**: Values are expressed as the median (Q1; Q3) and absolute count (percentage). **Abbreviations**: PWH, people with HIV; COVID-19, coronavirus disease 2019; SARS-CoV-2, severe acute respiratory syndrome coronavirus 2; ART, antiretroviral treatment; PWH, people with HIV; COVID-19, coronavirus disease 2019.

| Characteristics | Data |
| --- | --- |
| No. | 19 |
| Age (years) | 41 (33; 5) |
| Sex (male) | 15 (78.9%) |
| Previous SARS-CoV-2 Infection (+) | 6 (31.6%) |
| HIV infection |  |
| ART | 19 (100.0%) |
| CD4<sup>+</sup> count (cells/mm<sup>3</sup>) | 232 (158; 278) |
| CD4<sup>+</sup> count <200 cells/mm<sup>3</sup> | 6 (31.6%) |
| HIV viral load >50 copies/mL | 2 (10.5%) |

### Humoral response to the COVID-19 vaccine

GMFR values of the humoral response (anti-SARS-CoV-2 S IgG and inhibition of ACE2-S interaction) were higher than 8 after the second dose and higher than 4 after the booster dose for both the B.1 lineage and the Omicron variant (**Table 2**). Additionally, there was a strong correlation between anti-SARS-CoV-2 S IgG titers and ACE2-S interaction inhibition in all cases (**Supplementary Figure 1**).

**Table 2.** Humoral response in PWH generated by COVID-19 vaccine stratified by SARS-CoV-2 variants. **Statistics:** Values are expressed as geometric mean fold rise (95% confidence interval). Data were calculated using generalized linear mixed models adjusted (GLMM, see Statistical analysis section). **Abbreviations:** GMT, geometric mean titer; GMFR, geometric mean fold rise from baseline, 95% CI,IZ95% confidence interval; IgG, immunoglobulin G; ACE2, angiotensin-converting enzyme 2; S, Spike glycoprotein.

|  | Baseline | After the second dose |  | After the booster dose |  |
| --- | --- | --- | --- | --- | --- |
|  | GMT (95%CI) | GMT (95%CI) | GMFR (95%CI) | GMT (95%CI) | GMFR (95%CI) |
| **Wuhan (B.1)** |  |  |  |  |  |
| IgG antibody titers | 0.7 (0.2; 2.6) | 1866.5 (975.4; 3571.7) | 30.4 (18.7; 49.6) | 1134.9 (638.1; 2018.4) | 5 (3.5; 7.1) |
| Inhibition ACE2-S titer | 0.3 (0.1; 1.5) | 1326.5 (159.6; 11027.6) | 35.1 (14.7; 83.9) | 3780.1 (1395.5; 10239.6) | 7.6 (5.5; 10.5) |
| **Omicron** |  |  |  |  |  |
| IgG antibody titers | 0.3 (0.1; 0.8) | 539.5 (262.3; 1109.6) | 26 (18.4; 36.8) | 597.3 (319.3; 1117.4) | 5.3 (4.1; 6.7) |
| Inhibition ACE2-S titer | 0.1 (0.1; 0.1) | 28 (3; 264.8) | 11.6 (5.2; 25.9) | 234.3 (18.9; 2906.4) | 5.4 (3.7; 8) |

### Humoral response to COVID-19 vaccine between study groups

We found similar humoral responses (anti-SARS-CoV-2 S IgG and inhibition of ACE2-S interaction) to the B.1 lineage and Omicron variant between PWH and HC groups after the second and booster doses of the COVID-19 vaccine (q-value >0.05; **Supplementary Figure 2A & 2B**). Therefore, the humoral response in the PWH group was comparable to that of the HC group.

### Humoral response to COVID-19 vaccine between SARS-CoV-2 variants

After the second dose, both PWH and HC groups showed lower humoral responses against the Omicron variant compared to the B.1 lineage (q-value <0.05; **Figure 1A**). However, after the booster dose, we only found significant differences in ACE2-S interaction inhibition between the B.1 lineage and the Omicron variant among PWH (q-value =0.035; **Figure 1B**). The rate of PWH non-responders in the ACE2-S interaction inhibition (AUC = 0) against Omicron was significantly higher than against B.1 after the second dose (p-value = 0.007), but this difference was not significant after the booster dose (**Figure 2**). Therefore, the COVID-19 vaccine generated a significantly weaker humoral response against the Omicron variant than the B.1 lineage in both the HC and PWH groups. However, this response improved after the booster dose, although ACE2-S interaction inhibition in PWH remained significantly weaker.

**Figure 1.**
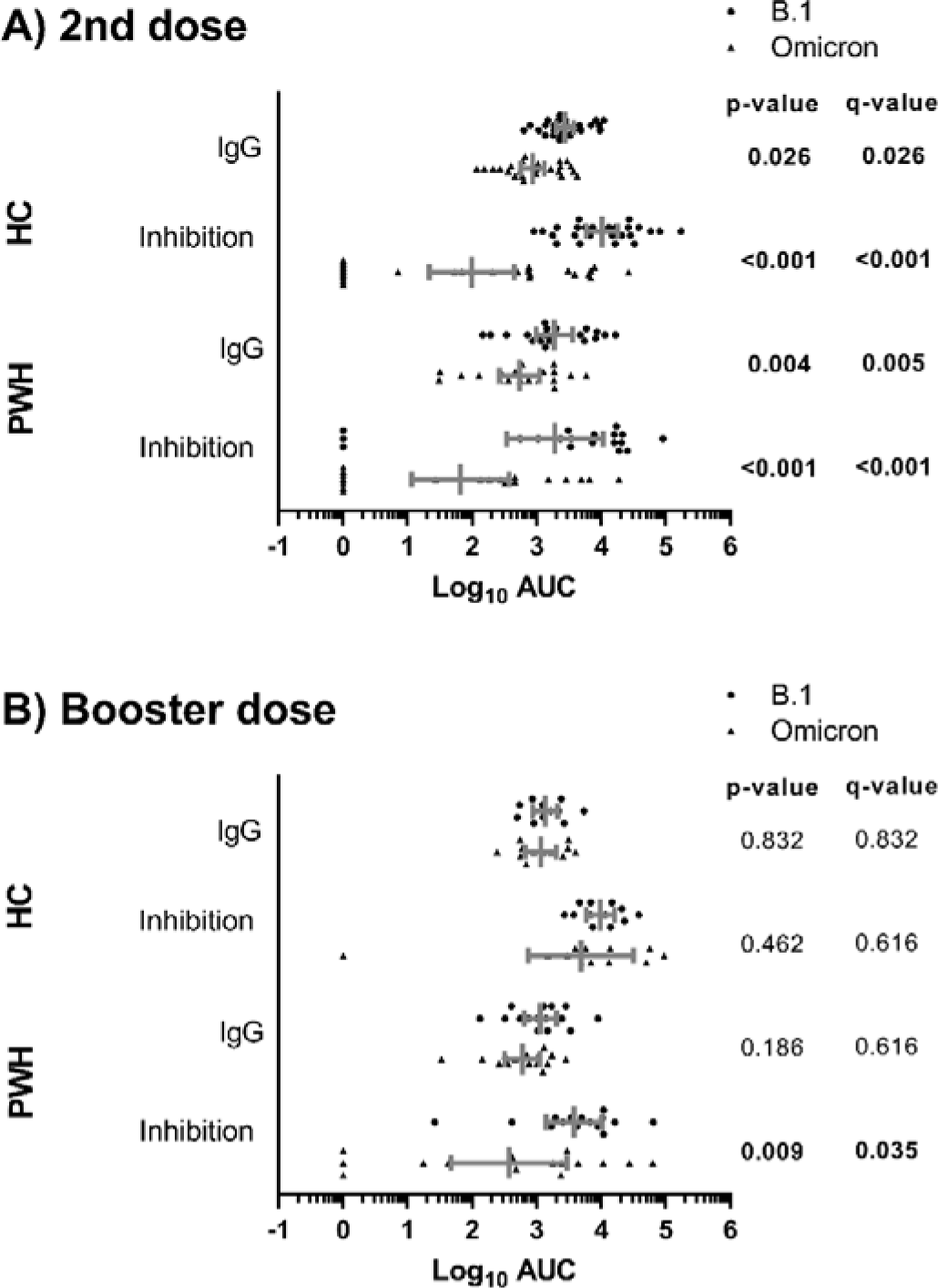
Comparison of humoral response to COVID-19 vaccine (anti-SARS-CoV-2 S IgG and inhibition of ACE2-S interaction) between SARS-CoV-2 variants in study groups after the second (A) and booster dose (B) of the COVID-19 vaccine. **Statistics:** The graph shows the geometric means and 95% confidence intervals in gray lines. P-values were calculated using generalized linear mixed models. Significant differences are shown in bold. **Abbreviations:** HC, healthy controls; PWH, people with HIV; Log10, base-10 logarithm; AUC, the area under the Curve; IgG, immunoglobulin G.

**Figure 2.**
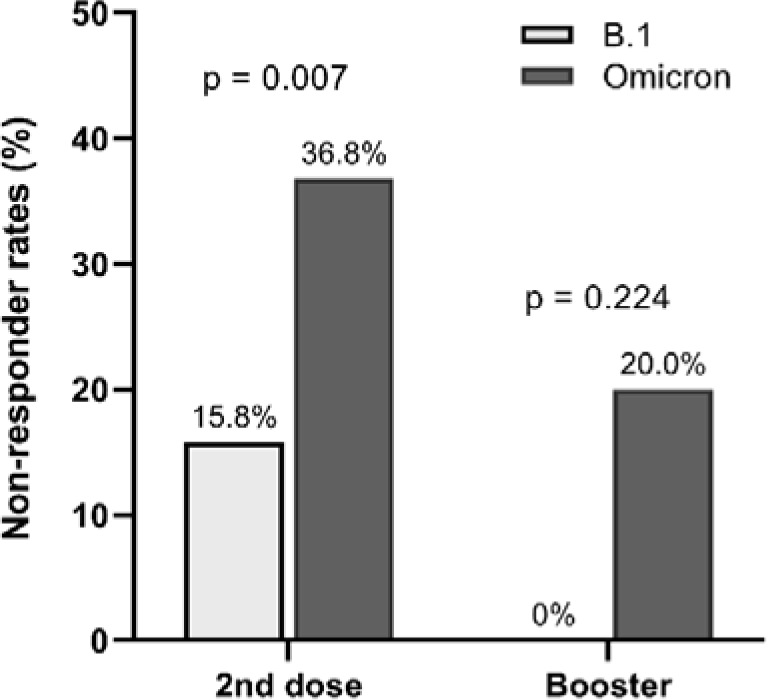
Rates of PWH non-responders in the inhibition of the ACE2-S interaction (AUC = 0), according to SARS-CoV-2 variants, after the second dose and the booster dose of the COVID-19 vaccine. **Statistics:** P-values were calculated using the Chi-square test.

## Discussion

PWH might have a weaker humoral response to vaccination, even with antiretroviral therapy. Moreover, although COVID-19 vaccines are effective against the original strain of SARS-CoV-2, the Omicron variant has impacted the efficacy of the immune response to vaccines. This study examined the humoral response to both the B.1 lineage and the Omicron variant following COVID-19 vaccination in PWH and HIV-negative controls. The major findings were: 1) PWH showed a strong humoral response after the second and booster doses. 2) The humoral response against the B.1 lineage and the Omicron variant after the second and booster doses was similar in the PWH and HC groups. 3) The humoral response to the Omicron variant was weaker than to the B.1 lineage in both groups but improved after the booster dose.

This study provides evidence of a robust humoral response against the COVID-19 vaccine in PWH. It shows a substantial increase in antibody levels against the B.1 lineage and the Omicron variant. The GMFR values were ≥8 after the second dose and ≥4 after the booster dose, indicating strong immunogenicity. These results align with previous studies showing a strong humoral response after the second and third doses of the COVID-19 vaccine [17]. Our data underscore the potential benefits of COVID-19 vaccination in protecting PWH against SARS-CoV-2 infection.

Moreover, anti-SARS-CoV-2 S IgG levels and the ability of these antibodies to inhibit the ACE2-S interaction were similar between PWH and the HC group for both the B.1 lineage and the Omicron variant, following the second and booster dose of the COVID-19 vaccine. These findings are consistent with previous research, which has also reported a comparable humoral immune response between PWH and HIV-negative controls following mRNA COVID-19 vaccination [18–20]. Of note, in these studies, the PWH were on ART with suppressed HIV viral loads and exhibited higher baseline CD4 counts compared to studies reporting diminished IgG responses in PWH relative to the HC group [20, 21]. In our study, patients were on suppressive ART with a median CD4 count <300 cells/mm^3^ and 31.6% had a CD4+ count <200 cells/mm^3^. Despite this, our study also found a robust immune response to the COVID-19 vaccine in PWH.

Our findings also showed that after the second vaccine dose, IgG levels against the Omicron variant and the inhibition of the ACE2-S interaction were significantly lower compared to those against the B.1 lineage. The Omicron variant contains many mutations, mainly within the S protein, the principal target of COVID-19 vaccines. These mutations help the virus evade the humoral response, reducing vaccine efficacy [22]. However, the response to Omicron significantly improved in both groups following administration of a booster dose. Significant differences between the B.1 lineage and the Omicron variant were observed only in the inhibition of the ACE2-S interaction in PWH after the booster. Despite this difference, the rate of PWH non-responders in ACE2-S interaction inhibition (AUC = 0) was not significantly different between the B.1 lineage and the Omicron variant after the booster dose. Consistent with these observations, previous studies have shown that despite the immune evasion of the Omicron variant to COVID-19 vaccines, booster doses are critical to enhance the efficacy of the immune response [23, 24]. These results underscore the crucial role of booster shots in sustaining protection against Omicron despite its partial evasion of the immune response elicited by the initial vaccination series.

### Study limitations

This study has several limitations that should be considered when interpreting the results. First, the sample size is limited, which reduces statistical power and increases the risk of false positive results. Second, the prospective nature of the study may have introduced biases, such as the loss of follow-up for some participants. Third, patient subgroup analysis, such as CD4 <200 or hybrid COVID-19 immunity, was not analyzed in detail due to the low number of patients. Fourth, our findings are primarily relevant to PWH who are on suppressive ART and may not apply to PWH who are not on ART with severe immune deficiency.

## Conclusions

PWH generated a strong humoral response to the COVID-19 vaccine against both the B.1 lineage and the Omicron variant, which were similar to that of individuals without HIV infection. However, our data suggest the need for booster doses to improve immunogenicity and update COVID-19 vaccines to new variants such as Omicron.

## List of abbreviations

Severe Acute Respiratory Syndrome Coronavirus 2 (SARS-CoV-2)

Coronavirus Disease 2019 (COVID-19)

People With Human Immunodeficiency Virus (PWH)

Antiretroviral therapy (ART)

Spike glycoprotein (S)

Angiotensin-converting enzyme 2 (ACE2)

Hospital Universitario Infanta Leonor (HUIL)

Healthy controls (HC)

Immunoglobulin G (IgG)

Immunoglobulin M (IgM)

Immunoglobulin A (IgA)

Enzyme-linked immunosorbent assay (ELISA)

Anti-SARS-CoV-2 S IgG (IgG against the SARS-CoV-2 spike protein)

Area under the curve (AUC)

Generalized Linear Mixed Models (GLMM)

Geometric mean fold rise (GMFR)

95% confidence interval (95%CI)

Geometric mean titer (GMT)

## Declarations

### Ethics approval and consent to participate

The HUIL Ethics Committee authorized the study (Ref.: 030-21), which was conducted following the Declaration of Helsinki. All participants gave their informed consent before enrollment.

### Consent for publication

Not applicable

### Availability of data and materials

The datasets used and/or analyzed during the current study are available from the corresponding author upon reasonable request.

### Competing interests

The authors have no competing interests, and funding sources played no role in the study’s design and research.

### Funding

The Centro de Investigación Biomédica en Red (CIBER) de Enfermedades Infecciosas (CB21/13/00044) funded the study.

## Data Availability

All data produced in the present study are available upon reasonable request to the authors

## Acknowledgments

The authors appreciate the collaboration of all the patients, medical and nursing staff and data managers who participated in the project. Besides, Dr. Jason McLellan (University of Texas, Austin, USA) generously provided the plasmid pαH, which encodes the S protein ectodomain (residues 1-1208) of SARS-CoV-2 2019-nCOV (GenBank: MN908947), stabilized in the prefusion conformation.

## Author contributions

Data curation: MJMG, PR, MQD, MMV, GC, JV, EJG, NB, AMG, GR, VM, MV, and JTM.

Investigation: MJMG, PR, MQD, MMV, and IM.

Data analysis and interpretation: MJMG, SR, and IM.

Supervision and visualization: PR, SR, and IM.

Funding acquisition: SR.

Drafting the article: MJMG, SR, and IM.

Critical revision of the article: PR.

All authors have read and approved the final manuscript.

## Authors’ information

Not applicable.

## Notes

### Competing Interest Statement

The authors have declared no competing interest.

### Funding Statement

This study was funded by The Centro de Investigacion Biomedica en Red (CIBER) de Enfermedades Infecciosas (CB21/13/00044).

### Author Declarations

The Ethics Committee of the Hospital Universitario Infanta Leonor (HUIL) gave ethical approval for this work (Ref.: 030-21)

