## Supplementary File for "Antibody response against SARS-CoV-2 spike protein in people with HIV after COVID-19 vaccination"

**Supplementary Figure 1.** Correlation between plasma IgG antibody levels against SARS-CoV-2 S protein (B.1 lineage and Omicron variant) and its capacity to inhibit the ACE2 receptor-S protein interaction in healthy controls and people with HIV after the second dose (A) and booster dose (B).

**Statistics**: Correlation analysis was performed using the Spearman test.

**Abbreviations**: AUC, the area under the curve; IgG, anti-SARS-CoV-2 S IgG; ACE2, angiotensin-converting enzyme 2; HC, healthy controls; PWH, people with HIV.

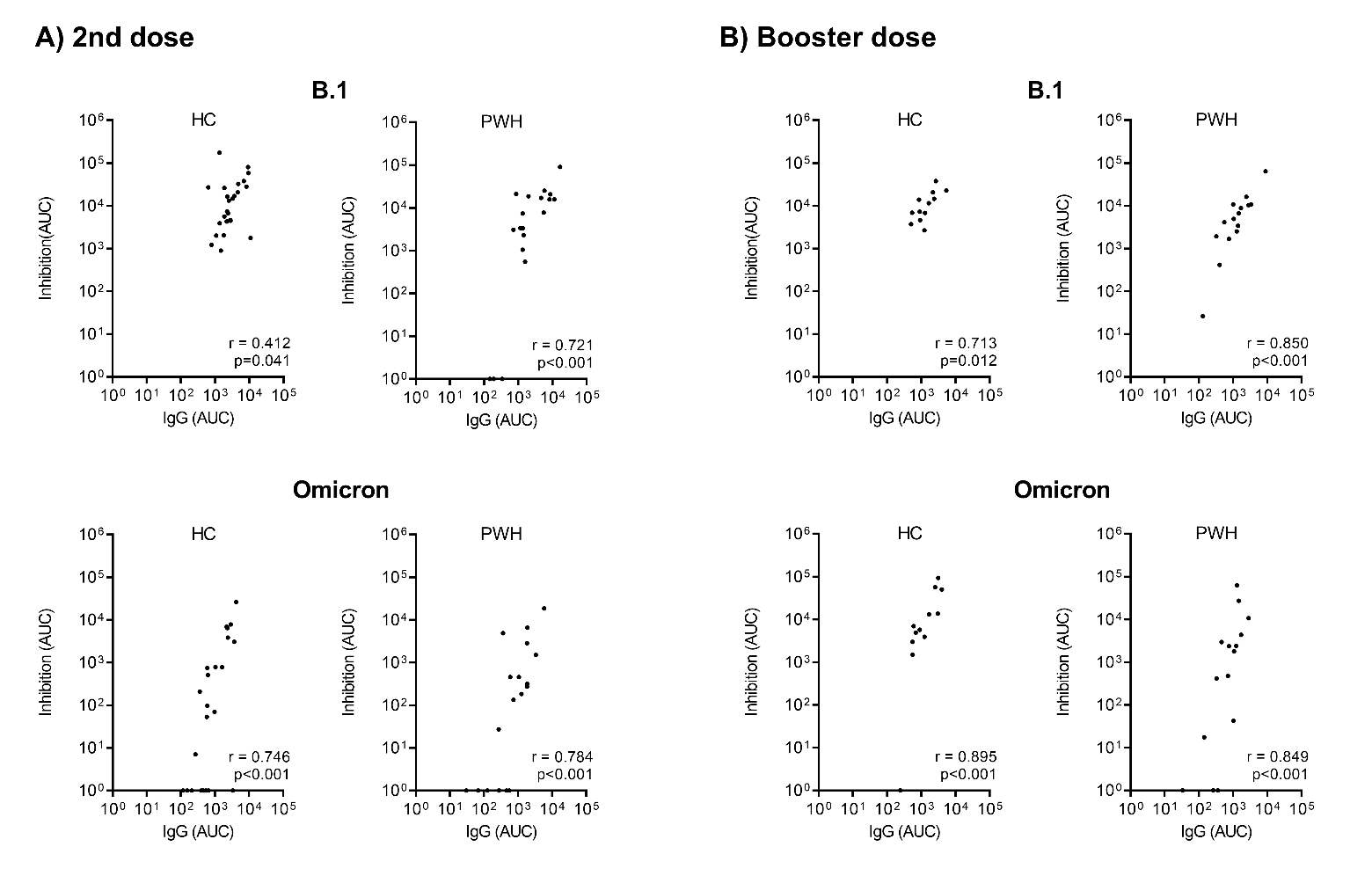

**Supplementary** **Figure 2.** Plasma IgG antibody levels against the SARS-CoV-2 S protein and ACE2-S inhibition titers, stratified by study groups and SARS-CoV-2 variants, weeks after the administration of the second (A) and booster dose (B) of the COVID-19 vaccine.

**Statistics**: The graph shows the geometric means and 95% confidence intervals in gray lines. P-values were calculated using generalized linear mixed models. Significant differences are shown in bold.

**Abbreviations**: HC, healthy controls; PWH, people with HIV; Log10, base-10 logarithm; AUC, the area under the Curve; IgG, immunoglobulin G.

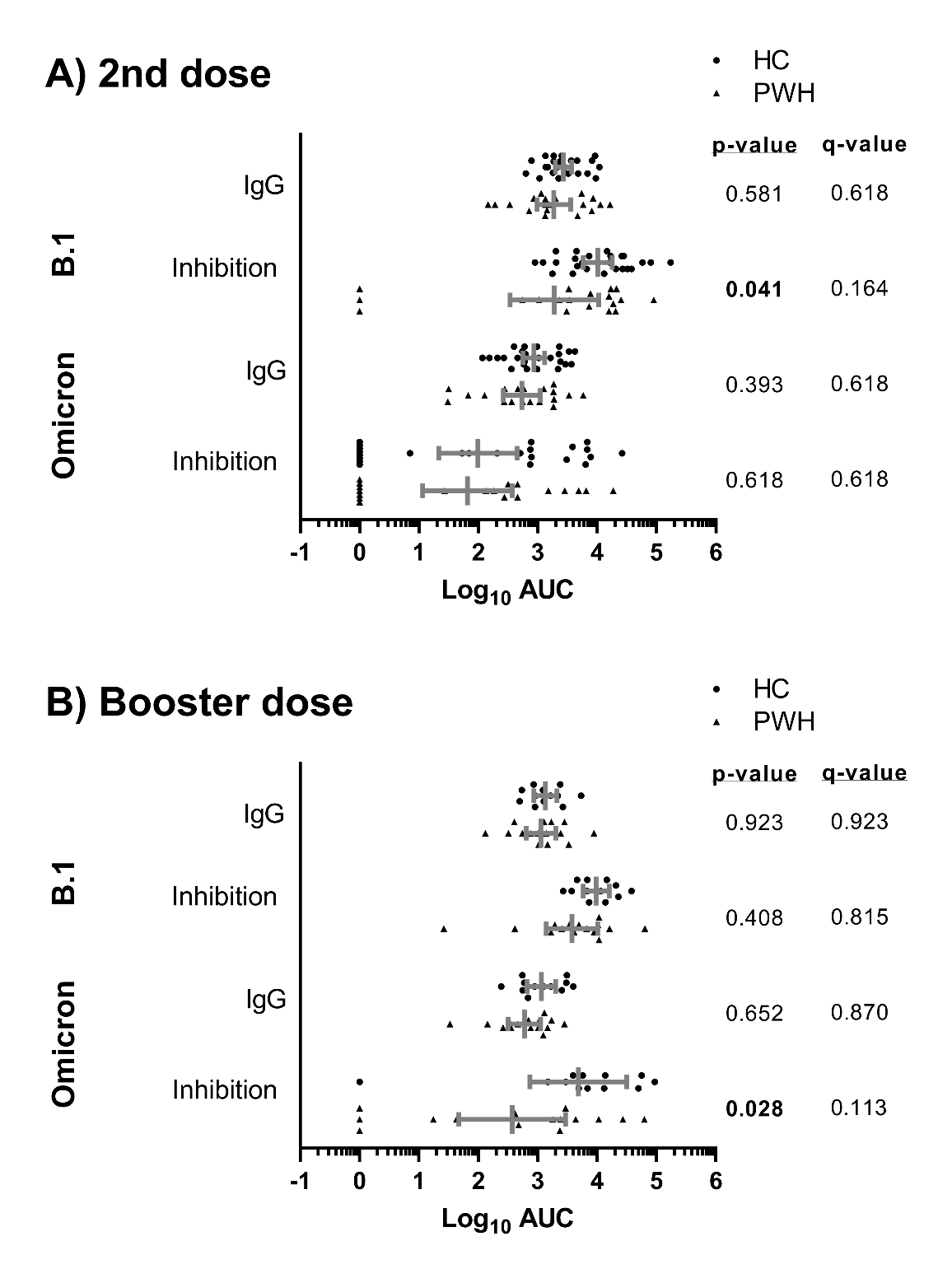
